# Relationship between Malnutrition and Functional Disability in Selected Community-Dwelling Geriatric Population in Bangladesh

**DOI:** 10.1101/2020.08.02.20167049

**Authors:** Mohammad Rahanur Alam, Md. Shahadat Hossain, Akibul Islam Chowdhury, Marufa Akhter, Abdullah Al Mamun, Sompa Reza

## Abstract

**Background:** The average life expectancy of the Bangladeshi population has been rising over the last decade due to the economic growth along with improved medicare. Although the increased number of geriatric people and their health is a matter of great concern, this issue remains unnoticed here.

**Objectives:** To assess the nutritional status of the functionality and to analyze the association between nutritional status and functional ability of the selected Bangladeshi geriatric population.

**Methods:** A community-based cross-sectional study was conducted among 400 participants, covering Chittagong, Noakhali, Comilla, and Jessore district of Bangladesh from December 2019 to February 2020. A standard and pretested questionnaire containing Mini Nutritional Assessment (MNA), Tinetti Performance Oriented Mobility Assessment (POMA), Activities of daily living scale (ADL), Lawton-Brody Instrumental Activities of Daily Living Scale (IADL), was used.

**Results:** According to our study, The prevalence of malnutrition and people at risk of malnutrition have been 25.4% and 58.8%, respectively. In the case of functionality, 63.3% of subjects have high falling risk, and 61.8% of subjects can independently do their daily activities while 38.3% are dependent. Furthermore, almost 80% of people are dependent in terms of doing living skills. High risk of falling (OR=10.823; 95% CI: 5.846-20.37; p<0.001), poor skill in doing ADL (OR=6.206; 95% CI: 4.021-9.581; p<0.001), along with dependency in performing IADL (OR=4.477; 95% CI: 2.833-7.075; p<0.001) are significantly associated with malnutrition.

**Conclusions:** Geriatric malnutrition can accelerate disability conditions, which can lead to early functional aging and subsequent loss in the quality of life.

## Introduction

The increase in the number of geriatric population has become a global phenomenon over the last decade. The number of older adults aged from 65 years and above is about 380 billion, and the number is projected to reach almost 2 billion by 2050 ^[1]^, and the number is higher for most high-income countries. Although the percentage of older people is higher in more developed countries, the pace of aging is rapid in developing countries ^[2]^. The number of elderly populations of Bangladesh is anticipated to be 16.2 and 42.2 million by 2025 and 2050, respectively, and it will represent little over 9% and 20% of the total population, respectively ^[3]^.

Now with the increase of the number of the aging population, a burning question arises: Are we ensuring proper health for this growing segment of the population? The significant challenges of elderly care in Bangladesh are the absence of any social security system, lack of employment opportunities, lack of financial source, and inadequate social support program ^[4].^

A disability can be referred to as any physical or psychological condition that leads to activity limitation and participation restrictions ^[5]^. During the aging process, several functional limitations occur due to physiological changes and chronic disease, as well as an impoverished lifestyle ^[6]^. Falls is very common among older due to several factors, and it constitutes a severe health problem. It is estimated that in one year, every third person over 65 years and every second over 85 years is subjected to falling ^[7]^. There be both intrinsic and extrinsic factors are associated with high-risk fall. These are deficits in sensory, arthritis, cognitive, central integrative, musculoskeletal abilities, decreased joint flexibility, the decline in vestibular function and dementia, etc., ^[8-11]^. Balance a complex process involves reception and integration of sensory inputs and execution of movements to control the center of gravity on the support base ^[12]^. Quality of life and functioning of older people mostly depend on gait and cognitive control. Older people who are poor functioning walk slower, have increased stride variability, and more mediocre performance on mobility tasks. Memory is vital in controlling balance and gait ^[13]^. During the last stage of the lifespan, older people go through physiological changes that decrease functional capacity and can be demonstrated as disabilities in the Basic Activities of Daily Living (ADL) and Instrumental Activities of Daily Living (IADL). ADL includes the basic everyday errands (e.g., eating, toileting, bathing) that are necessary to sustain healthy daily life ^[14]^. IADL encompasses complex activities such as food preparation, grocery shopping, transportation, and many more ^[15]^. The ability of individuals to carry out ADL and IADL is a measurement of functional ability in the elderly. In severe disabilities, there is a greater dependency on others for ADL and IADL. Disability increases illness and illness causes further disability and dependence on others ^[16]^. Malnutrition is a physiological condition characterized by either deficiency or excess of energy, protein, fat, and minerals ^[17]^. Nutritional status is associated with various biological, psychological, and socio-cultural factors ^[18]^. Geriatric malnutrition is often poorly diagnosed, which, along with reduced functional capacity, can increase the rate of morbidity and mortality ^[19]^. The present study aimed to assess the association between malnutrition and functional dependency in the selected population.

## Subjects and Methods

### Study design and participants

It was a community-based cross-sectional study. This study was carried from December 2019 to March 2020. The sample size was calculated by Epi Info software (version 7.2.3). The minimum sample size at a 95% confidence level was 348. We collected 400 samples from four different districts by a multistage sampling technique. At first, four districts (Chattogram, Comilla, Jashore, and Noakhali district) out of sixty-four districts were selected by convenient sampling (Figure 1a). Then, participants were chosen using a simple random sampling technique. People who were aged from 65 years or above and willingly agreed, participated in the study. The respondents who were mentally retarded as well as unable to complete the assessment were excluded from the study (Figure 1b). The study was conducted according to the declaration of Helsinki, and ethical approval was obtained from the institutional Ethical Committee. Informed consent from all participants was taken before the interview.

**Figure 1:**
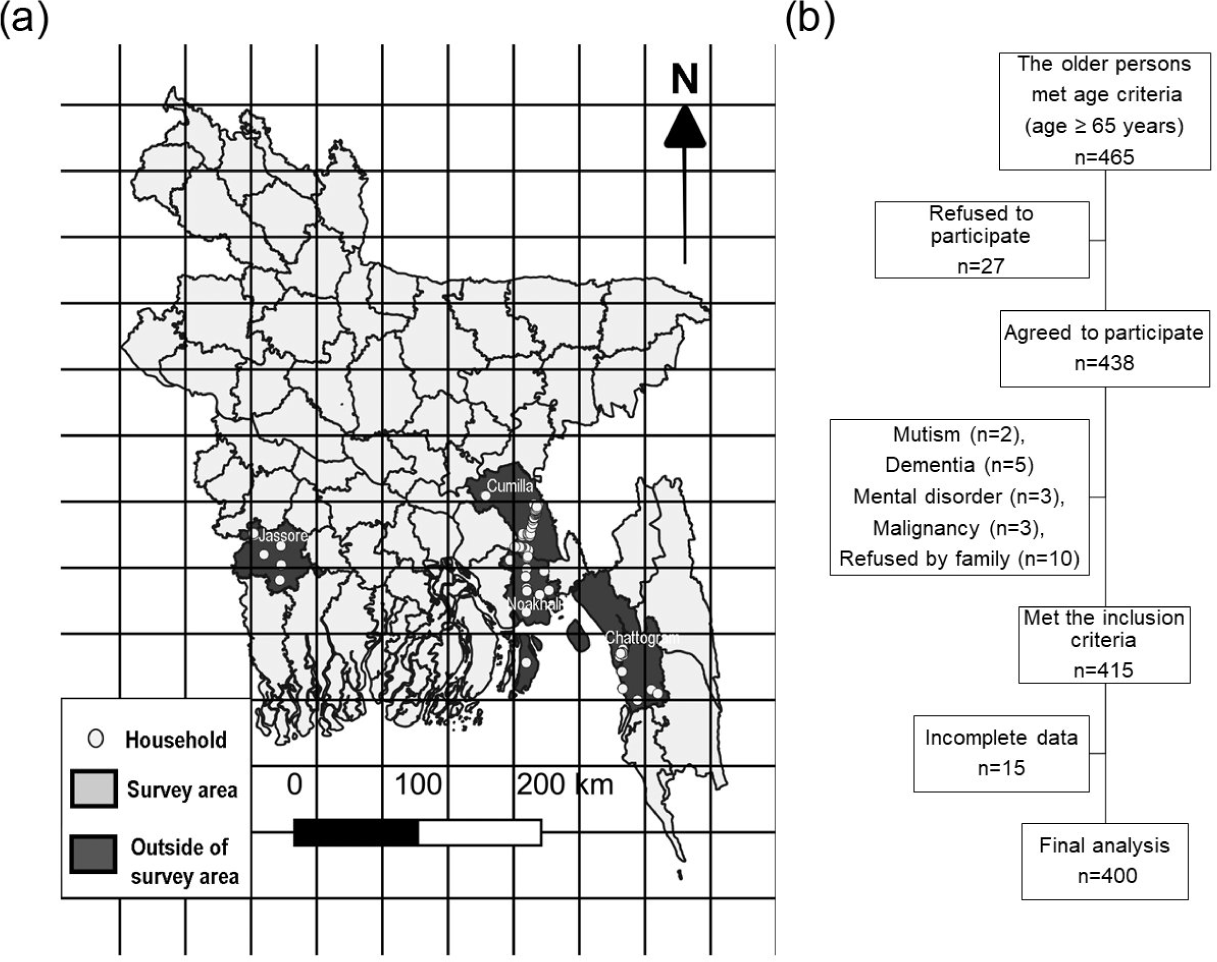
(a) Survey area (dark color indicates the survey area and circles represent selected households. The map was created by QGIS software, version 3.4.11), (b) The schematic diagram of study population selection.

### Questionnaire and data collection method

Participants were interviewed face to face using a pretested questionnaire in the local language. Android-based KoBoCollect (version 1.23.3) software was used to collect data from the participants. Three methods were employed to capture different types of functional disabilities, such as ADL limitations, IADL limitations, and POMA limitations. Self-reported Tinetti Performance Oriented Mobility Assessment (POMA) was used to measure the gait and balance ability as well as mobility. The total POMA scale consists of a balance scale and gait scale to perform balance maneuvers such as sitting, moving from a sitting position to a standing position, standing with eyes closed, and turning 360 degrees. Participants were assigned into three different categories according to the total score as “high fall risk” (<19 points), “medium fall risk” (20-24 points), or “minimal fall risk” (25-28 points) [20]. Activities of daily living (ADL) were evaluated using the Katz’ index. In the questionnaire, there were six questions on vital activities of daily life. A full score of six was defined as “good,” a score of 0-5 defined as “poor” ^[14]^. Lawton-Brody instrumental activities of the daily living (IADL) scale was used to evaluate an individual’s capacity to participate in more complex activities, which are necessary for functioning in community settings by assessing the following eight areas of occupational performance: the ability to use a telephone, shopping, food preparation, housekeeping, laundry, mode of transportation, responsibility for own medications, and ability to handle finances. On a scale of eight, a score of eight was classified as independent, and between 0 to 7 was indicated as dependent ^[15]^. Mini Nutritional Assessment (MNA) was used to assess geriatric nutritional status. With a maximum score of 30, a score of 24 or higher indicated normal nutritional status, 19 to 23 indicated at risk of malnutrition, and 18 or lower represented malnutrition ^[21]^.

### Statistical Analysis

The Categorical variables are presented as frequency and percentages, where continuous variables are presented as mean and standard deviation. A Chi-square test was used to determine the correlation between categorical variables such as age, sex, and nutritional status. One way ANOVA was performed among the three nutritional status groups for continuous variables such as POMA score, Lawton-Brody IADL score, and Katz’ index. Data were checked for homoscedasticity by Levene’s test. Welch ANOVA was performed in the cases of deviation from homogeneity. Multinomial regression analysis was done to determine the association between nutritional status and functionality. The level of significance for the statistical tests was set at 0.05. Data analysis was done using Statistical Package for Social Sciences (SPSS) Version 23.0.

## Results

The mean age of the study population is 72.1 ± 7.0 years. Over three-quarters of the participants were classified as elderly (65 through 74 years), About 22% were old (76 through 90 years), and the rest were very old (over 91 years old).

According to the MNA score, a quarter portion of the sample population has been observed to be malnourished, and more than half of the participants are at the risk of malnutrition. While measuring gait and balance tests, it has been found that more than 60% of the participants were at risk of high falling, and the percentage of medium and minimal falling risk is about 20.3% and 16.4%, respectively. According to the Katz’ index of Independence in Activities of Daily Living (ADL), 38.3% of people are dependent on doing daily activities. Assessment of independent living skills by The Lawton-Brody Instrumental Activities of Daily Living (IADL) Scale has shown that almost 80% of the geriatric population is dependent in terms of doing living skills (Table 1).

**Table 1:** Sociodemographic characteristics, nutritional status, and functionality of the study population.

| Variable | Category | Frequency | Percentage |
| --- | --- | --- | --- |
| **Age** | Elderly | 304 | 76.0 |
|  | Old | 87 | 21.70 |
|  | Very Old | 9 | 2.30 |
| **Sex** | Female | 200 | 50.0 |
|  | Male | 200 | 50.0 |
| **MNA classification** | Malnourished | 102 | 25.40 |
|  | At-risk | 235 | 58.80 |
|  | Normal | 63 | 15.80 |
| **POMA classification** | Minimal fall risk | 66 | 16.50 |
|  | Medium Fall Risk | 81 | 20.20 |
|  | High Fall Risk | 253 | 63.30 |
| **Katz' ADL classification** | Good | 247 | 61.70 |
|  | Poor | 153 | 38.30 |
| **Lawton-Brody IADL classification** | Independent | 81 | 20.30 |
|  | Dependent | 319 | 79.70 |

MNA: Mini Nutritional Assessment, POMA: Performance Oriented Mobility Assessment, ADL: Independence in Activities of Daily Living, IADL: Instrumental Activities of Daily Living

The correlation among MNA, POMA, Katz’ ADL, and Lawton-Brody IADL with sex and age category of the respondents has been presented in Table 2. A statistically significant correlation between nutritional status and the Lawton-Brody IADL scale with both age and gender categories of the respondents has been observed. Moreover, the correlation between the Katz’ ADL index and age category has also shown a significant result. However, no significant correlation between the POMA score and the Katz’ ADL in the case of gender has been found.

**Table 2:**
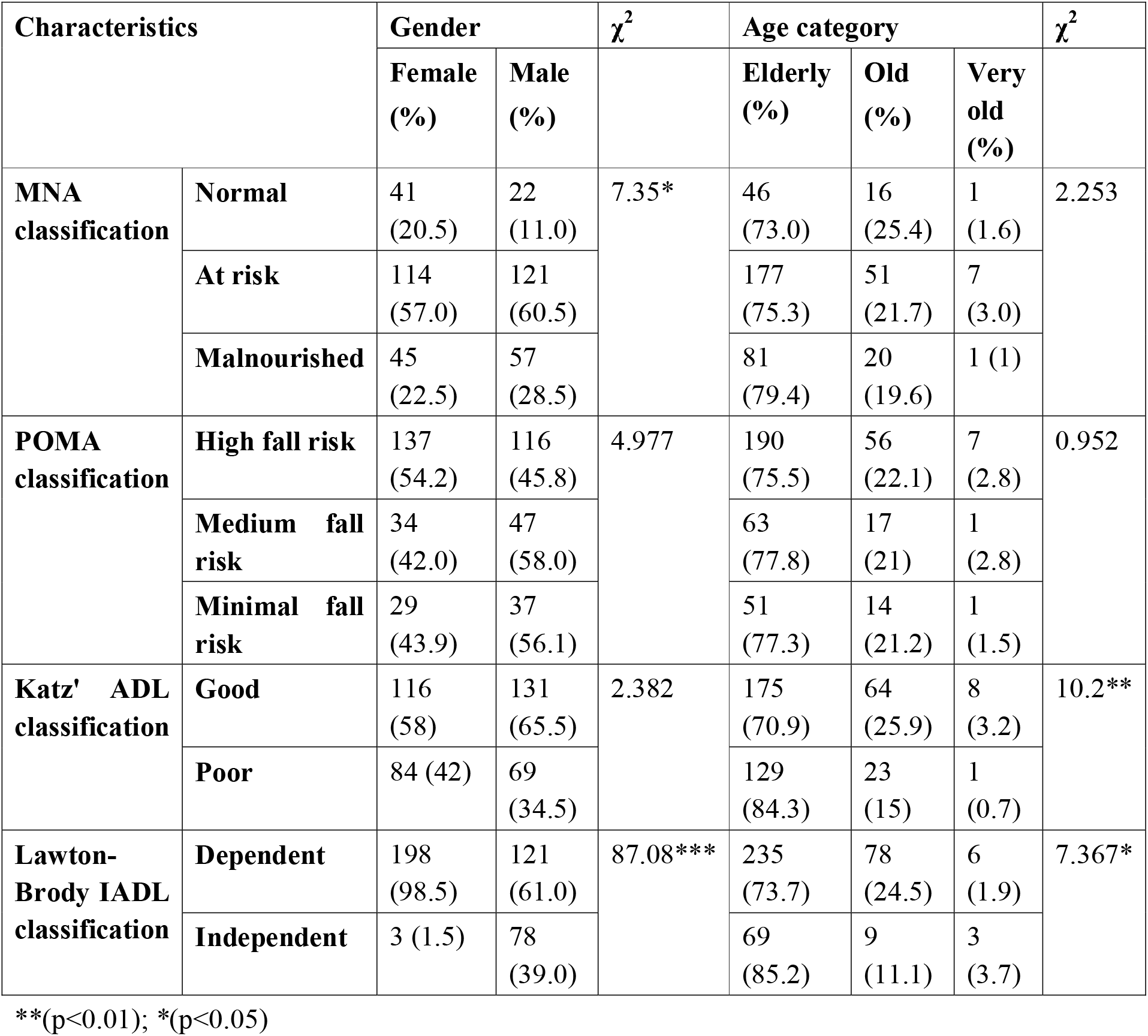
Correlation of Nutritional Status, POMA, Katz’ ADL, and Lawton-Brody category with gender and age category of the respondents.

| Characteristics | | Gender | | $\chi^2$ | Age category | | | $\chi^2$ |
| --- | --- | --- | --- | --- | --- | --- | --- | --- |
|  |  | Female (%) | Male (%) |  | Elderly (%) | Old (%) | Very old (%) |  |
| **MNA classification** | **Normal** | 41<br>(20.5) | 22<br>(11.0) | 7.35* | 46<br>(73.0) | 16<br>(25.4) | 1<br>(1.6) | 2.253 |
|  | **At risk** | 114<br>(57.0) | 121<br>(60.5) |  | 177<br>(75.3) | 51<br>(21.7) | 7<br>(3.0) |  |
|  | **Malnourished** | 45<br>(22.5) | 57<br>(28.5) |  | 81<br>(79.4) | 20<br>(19.6) | 1 (1) |  |
| **POMA classification** | **High fall risk** | 137<br>(54.2) | 116<br>(45.8) | 4.977 | 190<br>(75.5) | 56<br>(22.1) | 7<br>(2.8) | 0.952 |
|  | **Medium fall risk** | 34<br>(42.0) | 47<br>(58.0) |  | 63<br>(77.8) | 17<br>(21) | 1<br>(2.8) |  |
|  | **Minimal fall risk** | 29<br>(43.9) | 37<br>(56.1) |  | 51<br>(77.3) | 14<br>(21.2) | 1<br>(1.5) |  |
| **Katz' ADL classification** | **Good** | 116<br>(58) | 131<br>(65.5) | 2.382 | 175<br>(70.9) | 64<br>(25.9) | 8<br>(3.2) | 10.2** |
|  | **Poor** | 84 (42) | 69<br>(34.5) |  | 129<br>(84.3) | 23<br>(15) | 1<br>(0.7) |  |
| **Lawton-Brody IADL classification** | **Dependent** | 198<br>(98.5) | 121<br>(61.0) | 87.08*** | 235<br>(73.7) | 78<br>(24.5) | 6<br>(1.9) | 7.367* |
|  | **Independent** | 3 (1.5) | 78<br>(39.0) |  | 69<br>(85.2) | 9<br>(11.1) | 3<br>(3.7) |  |

\*\*( $p < 0.01$ ); \*( $p < 0.05$ )

All of the scores are significantly lower in the malnourished group and at-risk group compared to the normal group (Table 3).

**Table 3:** Subject characteristics according to nutritional status.

| Variable | Normal<br>n=63 | At-risk<br>n=235 | Malnourished<br>n=102 | Total<br>n=400 | F<br>Statistic | Sig |
| --- | --- | --- | --- | --- | --- | --- |
| | Mean ± SD | Mean ± SD | Mean ± SD | Mean ± SD | | |
| **POMA (points)** | 21.8 ± 4.6 | 16.4 ± | 8.4 ± 6.3 | 15.2 ± | 122.468<sup>†</sup> | .00*** |
|  |  | 5.8 |  | 7.3 |  |  |
| **Katz' ADL (points)** | $6.0 \pm 0.1$ | $5.5 \pm 0.8$ | $3.9 \pm 2.1$ | $5.2 \pm 1.5$ | $85.463^{\dagger}$ | .00*** |
| **Lawton-Brody IADL (points)** | $4.6 \pm 1.8$ | $3.9 \pm 1.8$ | $1.8. \pm 1.9$ | $3.5 \pm 2.1$ | 60.51 | .00*** |

<sup>†</sup>Asymptotically F distributed; therefore robust tests of equality of means were performed; \*\*\*( $p < 0.001$ )

Figure 2a depicts that the POMA score has been proportionally increased with nutritional status (MNA) score, which means that those who have a higher score of MNA score (normal nutritional status), will have a better score of Tinetti Performance Oriented Mobility Assessment (POMA) (minimal fall risk). Both Katz’ ADL and Lawton-Brody IADL score significantly correlate with the MNA score (Figure 2b, 2c).

**Figure 2:**
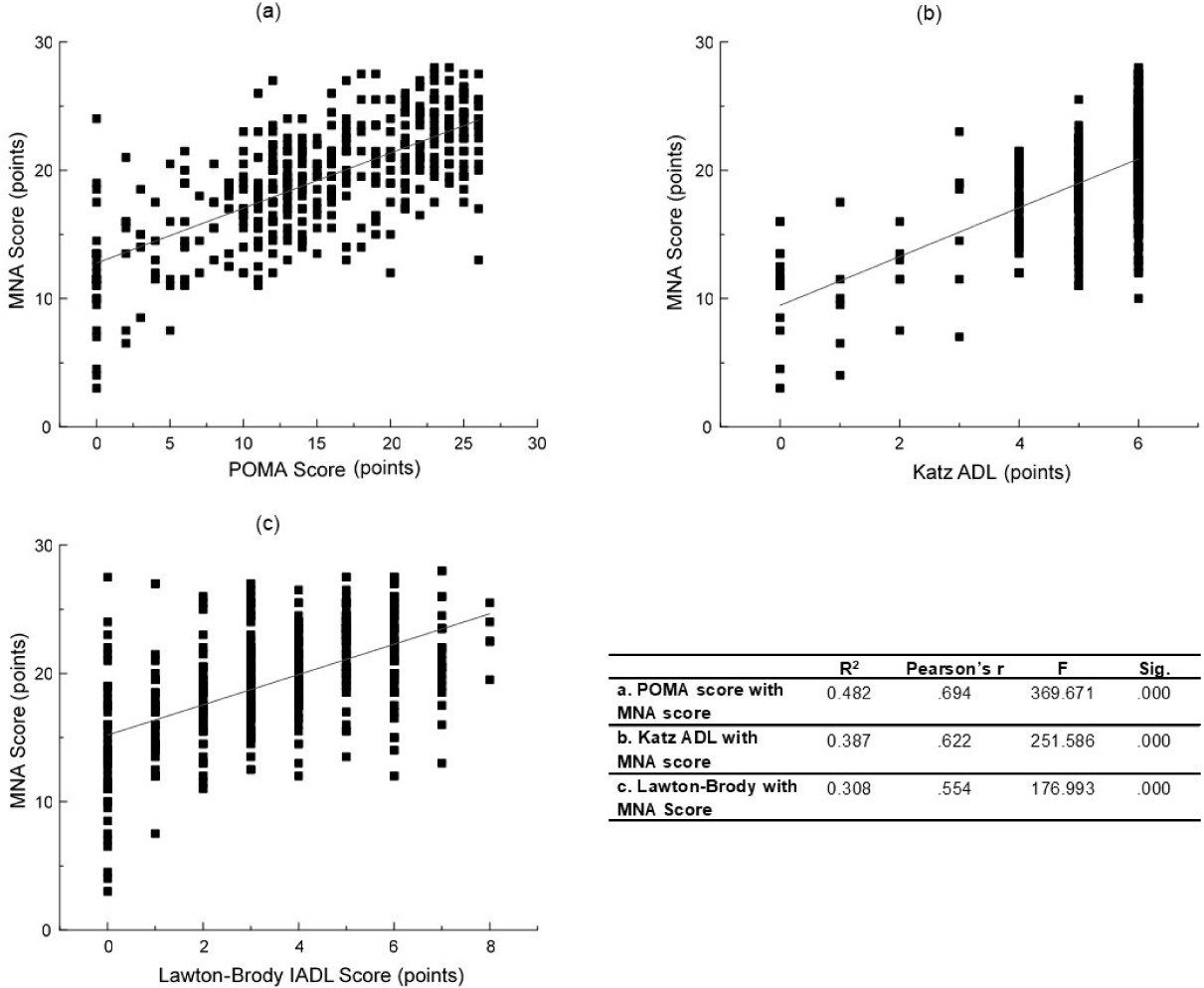
Relationship among MNA score and POMA score (a), Katz’ ADL (b) and Lawton-Brody IADL score (c)

The association of POMA, Katz’ ADL, and Lawton-Brody IADL with the Nutritional Status of the respondents has been presented in Table 4. The correlation among POMA, Katz’ ADL, and Lawton-Brody IADL with the Nutritional Status of the respondents show a statistically significant result. High-risk falls have been found to be significantly associated with nutritional status. The malnourished and people at risk of malnutrition have significantly high fall risk. Furthermore, if the person is malnourished, the risk of being dependent on others in engaging in necessary lifestyle activities as well as more complex functional activities for functioning in community settings is significantly higher (Table 4).

**Table 4:** Correlation and Regression analysis of POMA, Katz’ and Lawton-Brody category with the Nutritional Status of the respondents.

| Characteristics | | MNA classification | | | $\chi^2$ | OR<br>(95% CI) |
| --- | --- | --- | --- | --- | --- | --- |
|  |  | Normal (%) | At-risk (%) | Malnourished (%) |  |  |
| **POMA classification** | **High fall risk** | 11 (4.3) | 147 (58.1) | 95 (37.6) | 99.005*** | 10.823***<br>(5.846-20.37) |
|  | **Medium fall risk** | 25 (30.9) | 50 (61.7) | 6 (7.4) |  | 1.683<br>(0.931-3.041) |
|  | **Minimal fall risk** | 27 (40.9) | 38 (57.6) | 1 (1.5) |  | 1 (Ref) |
| **Katz' ADL classification** | **Good** | 62 (25.1) | 155 (62.8) | 30 (12.1) | 82.775*** | 1 (Ref) |
|  | **Poor** | 1 (0.7) | 80 (52.2) | 72 (47.1) |  | 6.206***<br>(4.021-9.581) |
| **Lawton-Brody IADL classification** | **Dependent** | 33 (10.3) | 187 (58.6) | 99 (31) | 48.149*** | 4.477***<br>(2.833-7.075) |
|  | **Independent** | 30 (37) | 48 (59.3) | 3 (3.7) |  | 1 (Ref) |

\*\*\*( $p < 0.001$ )
OR, Odds Ratio
CI, Confidence Interval

## Discussion

This study focuses on the relationship between nutritional status and functional health of the community living geriatric population in Bangladesh. The result demonstrates that more than 84% were either malnourished or at risk of malnutrition, which corresponds with a previous study conducted in Bangladesh ^[6]^. As there is no available gold standard technique of geriatric nutritional assessment ^[22]^, Mini nutritional assessment (MNA) was used to assess the nutritional status of the study population. Although MNA is not very reliable for assessing the nutritional status of hospitalized or nursing patients, it can provide reasonably accurate estimates for community living elderly than simple anthropometry ^[23, 24]^. Additionally, POMA, Katz’ ADL, Lawton IADL scales provided a non-invasive and rapid assessment of functional ability ^[25]^.

In the study, malnourished and people at the risk of malnutrition have shown significantly lower POMA scores. Therefore, people with high falling risk have a higher tendency of being malnourished (OR: 10.823, 95% CI:5.846-20.37, p<0.001), which is in line with the literature ^[10, 26, 27]^. Malnutrition can lead to significant loss of muscle mass ^[28]^, which is associated with impaired balance and high risk falling in older adults ^[29]^.

The overall independence rate is substantially lower because one may be independent in some functions, but in doing some other functions, they are dependent. For example, most of the male are dependent on food preparation; on the other hand, most of the female was dependent on someone to use telephone, shopping, transportations and handling finance on their own. It can be explained by the fact that traditionally, food preparation is considered as “women’s work” while working outside is considered as “men’s job” ^[30]^. This phenomenon explains the higher number of dependencies among women. Malnutrition is associated with Poor ADL (OR: 6.206, 95% CI: 4.021-9.581, p<0.001) as well as dependent on help to doing IADL (OR: 4.477, 95% CI: (2.833-7.075, p<0.001). Since both ADL and IADL include functions related to food intakes such as eating, food preparation, grocery shopping. Limitations in these domains can lead to inadequate food intake, eventually to malnutrition. With the increasing age, their dependency in terms of doing ADL and IADL also have increased significantly.

Our study had several limitations. First of all, our study was a questionnaire-based cross-sectional study. Therefore, the result can only provide a snapshot of the current nutritional status and functional health of the study population. Though poor functionality is associated with malnutrition, causation cannot be established due to the multifactorial nature of malnutrition. Secondly, MNA, Lawton IADL, and Katz’ ADL scale mainly based on self-reported information. There is maybe a possibility of overestimation or underestimation ^[25, 31]^. Finally, the elderly population also encounters with psychological symptoms due to malnutrition ^[32]^. Although the definition of disability includes psychological impairment, we did not consider psychological impairments in our study.

In conclusion, the prevalence of malnutrition among older people in Bangladesh deplorable and poor functional health has shown a significant association with geriatric malnutrition. Enhancing the nutritional status can decrease the risk of functional disability, thus improving quality of life. Necessary measures to improve functional health and nutrition conditions should be taken at both community and national levels, such as dissemination of knowledge on nutrition and physical exercise, engaging the elderly population in social activities.

## Data Availability

The datasets generated during this study are available from the corresponding author on a reasonable request.

## References

1. Nations, U., World Population Ageing 2017-Highlights. Department of Economic and Social Affairs, 2017.

2. Morley, J.E. and D.R. Thomas, Geriatric nutrition. 2007: CRC Press.

3. Haque, M.M., et al., Health and Nutritional Status of Aged People. Chattagram Maa-O-Shishu Hospital Medical College Journal, 2014. 13(3): p. 30–34.

4. Ferdousi, N., Protecting Elderly People in Bangladesh: An Overview. Jurnal Undang-undang dan Masyarakat, 2020. 24.

5. WHO, International classification of functioning, disability and health: ICF. 2001: World Health Organization.

6. Ferdous, T., et al., Nutritional status and self-reported and performance-based evaluation of physical function of elderly persons in rural Bangladesh. Scandinavian journal of public health, 2009. 37(5): p. 518–524.

7. Borowicz, A., et al., Assessing gait and balance impairment in elderly residents of nursing homes. Journal of physical therapy science, 2016. 28(9): p. 2486–2490.

8. Shumway-Cook, A., et al., The effect of multidimensional exercises on balance, mobility, and fall risk in community-dwelling older adults. Physical therapy, 1997. 77(1): p. 46–57.

9. Teixeira-Leite, H. and A.C. Manhães, Association between functional alterations of senescence and senility and disorders of gait and balance. Clinics, 2012. 67(7): p. 719–729.

10. Mecagni, C., et al., Balance and ankle range of motion in community-dwelling women aged 64 to 87 years: a correlational study. Physical Therapy, 2000. 80(10): p. 1004–1011.

11. Owsley, C. and G. McGwin Jr, Association between visual attention and mobility in older adults. Journal of the American Geriatrics Society, 2004. 52(11): p. 1901–1906.

12. Karuka, A.H., J.A. Silva, and M.T. Navega, Analysis of agreement of assessment tools of body balance in the elderly. Brazilian Journal of Physical Therapy/Revista Brasileira de Fisioterapia, 2011. 15(6).

13. Iersel, M.B.v., et al., Executive functions are associated with gait and balance in community-living elderly people. The Journals of Gerontology Series A: Biological Sciences and Medical Sciences, 2008. 63(12): p. 1344–1349.

14. Katz, S., et al., Studies of illness in the aged: the index of ADL: a standardized measure of biological and psychosocial function. Jama, 1963. 185(12): p. 914–919.

15. Lawton, M.P. and E.M. Brody, Assessment of older people: self-maintaining and instrumental activities of daily living. The gerontologist, 1969. 9(3_Part_1): p. 179-186.

16. Cervantes Becerra, R.G., et al., Health status of the elderly in primary health care practices using an integral geriatric assessment. Atención Primaria, 2015. 47(6): p. 329–335.

17. Freyer, K., M.J.C. Nuijten, and J.M.G.A. Schols, The budget impact of oral nutritional supplements for disease related malnutrition in elderly in the community setting. Frontiers in pharmacology, 2012. 3: p. 78.

18. Bailly, N., I. Maître, and V. Van Wymelbeke, Relationships between nutritional status, depression and pleasure of eating in aging men and women. Archives of gerontology and geriatrics, 2015. 61(3): p. 330–336.

19. Harith, S., et al., The magnitude of malnutrition among hospitalized elderly patients in university Malaya medical centre. Health Environ J, 2010. 1(2): p. 64–72.

20. Tinetti, M.E., Performanceloriented assessment of mobility problems in elderly patients. Journal of the American Geriatrics Society, 1986. 34(2): p. 119–126.

21. Vellas, B., et al., The Mini Nutritional Assessment (MNA) and its use in grading the nutritional state of elderly patients. Nutrition, 1999. 15(2): p. 116–122.

22. Aziz, N.A.S.A., et al., Assessing the nutritional status of hospitalized elderly. Clinical interventions in aging, 2017. 12: p. 1615.

23. Hudgens, J. and B. Langkamp-Henken, The Mini Nutritional Assessment as an assessment tool in elders in long-term care. Nutr Clin Pract, 2004. 19(5): p. 463–70.

24. Hailemariam, H., P. Singh, and T. Fekadu, Evaluation of mini nutrition assessment (MNA) tool among community dwelling elderly in urban community of Hawassa city, Southern Ethiopia. BMC Nutrition, 2016. 2(1): p. 11.

25. Graf, C., The Lawton instrumental activities of daily living (IADL) scale. The gerontologist, 2009. 9(3): p. 179–186.

26. Chu, J.-J., et al., A poor performance in comprehensive geriatric assessment is associated with increased fall risk in elders with hypertension: a cross-sectional study. Journal of geriatric cardiology: JGC, 2015. 12(2): p. 113.

27. Thomas, J.I. and J.V. Lane, A pilot study to explore the predictive validity of 4 measures of falls risk in frail elderly patients. Archives of physical medicine and rehabilitation, 2005. 86(8): p. 1636–1640.

28. Pierik, V.D., et al., High risk of malnutrition is associated with low muscle mass in older hospitalized patients - a prospective cohort study. BMC geriatrics, 2017. 17(1): p. 118–118.

29. Szulc, P., et al., Low Skeletal Muscle Mass Is Associated With Poor Structural Parameters of Bone and Impaired Balance in Elderly Men—The MINOS Study. Journal of Bone and Mineral Research, 2005. 20(5): p. 721–729.

30. Cain, M., S.R. Khanam, and S. Nahar, Class, Patriarchy, and Women’s Work in Bangladesh. Population and Development Review, 1979. 5(3): p. 405–438.

31. Reijneveld, S.A., J. Spijker, and H. Dijkshoorn, Katz’ADL index assessed functional performance of Turkish, Moroccan, and Dutch elderly. Journal of clinical epidemiology, 2007. 60(4): p. 382–388.

32. Kvamme, J.-M., et al., Risk of malnutrition is associated with mental health symptoms in community living elderly men and women: The Tromsø Study. BMC psychiatry, 2011. 11(1): p. 112.

